# Type 1 Diabetes Genetic Risk Score classifies diabetes subtypes in Indians: Impact of HLA diversity on the lower discriminative ability

**DOI:** 10.1101/2024.08.31.24312873

**Authors:** Alagu Sankareswaran, Pooja Kunte, Diane P Fraser, Mobeen Shaik, Michael N Weedon, Richard A Oram, Chittaranjan S Yajnik, Giriraj R Chandak

## Abstract

**Objectives:** Genetic Risk scores (GRS) classify diabetes types, type 1 (T1D) and type 2 (T2D) in Europeans but the power is limited in other ancestries. We explored the performance of T1DGRS and potential reasons for inferior discrimination ability in diabetes-type classification in Indians.

**Research Design and Methods:** In a well-characterized Indian cohort comprising 645 clinically diagnosed T1D, 1153 T2D and 327 controls, we estimated the discriminative ability of T1DGRS (comprising 67 SNPs from Europeans) using receiver operating characteristics-area under the curve (ROC-AUC). We also compared the islet autoantibody status (AA), frequency and effect size of various HLA alleles/haplotypes between Indians and Europeans.

**Results:** The T1DGRS was discriminative of T1D from T2D and controls but the ability is lower in Indians than Europeans (AUC=0.83 vs 0.92 respectively, p<0.0001). The T1DGRS was higher in AA-positive patients compared to AA-negative patients [13.01 (12.79-13.23) vs 12.09 (11.64-12.56)], p<0.0001) and showed greater discrimination in the AA-positive T1D (ROC-AUC 0.85). While association of common HLA-DQA1∼HLA-DQB1 haplotypes with T1D is replicated, important differences in the risk allele frequency, nature/direction and magnitude of association between Indians and Europeans were noted.

**Conclusions:** A T1DGRS derived from Europeans is discriminative of T1D in Indians, highlighting similarity in heritability of T1D. Differences in allele frequency, effect size and directionality, especially in the HLA region are important contributors to inferior discrimination performance of T1DGRS in Indians. Further studies of diverse populations may improve its performance.

## Introduction

Diabetes is a major non-communicable disease (NCD) characterized by insulin resistance, deficient insulin secretion, or a combination of both. The two primary types of diabetes, Type 1 (T1D) and Type 2 (T2D), account for approximately 98% of diabetes cases worldwide^1^. While T1D commonly occurs in children, it can occur at any age^2,3,4^. In contrast, T2D is a lifestyle-related disorder that typically occurs later in life, driven by factors such as obesity and insulin resistance. In Indians, T2D manifests at lower body mass index (BMI) and an earlier age compared to individuals of European descent, hence BMI and late-onset are no longer distinctive features of T2D, particularly among Indians^5^. The overlapping clinical features between T1D and T2D pose challenges in their accurate diagnosis which is critical since the optimal treatments for these conditions differ significantly^6^. Misclassified T1D cases can lead to severe complications, such as Diabetic Ketoacidosis, due to insufficient insulin administration, and increased risk of chronic complications due to poor glycemic control, thus, an early and precise diagnosis of T1D is crucial for effective management and prevention of complications^7^. On the other hand, people with T2D commonly have a relative insulin deficiency in the context of insulin resistance and may require a combination of oral hypoglycemic drugs and insulin therapy for better glycaemic control^8^. Apart from the age of presentation and C-peptide, islet autoantibody (AA) testing has been employed to aid in the screening and diagnosis of T1D but is highly expensive^8–10^. Further, their levels keep fluctuating with the disease treatment and duration, hence, there is a need to develop novel tools for accurate diagnosis.

In recent years, genetic risk scores (GRS) calculated from multiple risk genetic variants have emerged as a promising tool for assessing an individual’s genetic risk and predicting disease susceptibility. The increasing popularity of polygenic scores stems from their potential applications in both screening and diagnosis. Both T1D and T2D have a strong hereditary component, but our knowledge regarding the genetic features of T1D in the Indian population is limited, as most genome-wide association studies (GWAS) have predominantly focused on individuals of European ancestry^11–13^. Further, the genetic architecture of the Indian population [South Asians (SAS)] differs from that of Europeans^14^. We have earlier provided evidence of genetic and mutational heterogeneity in other complex diseases in Indians^15,16^. On similar lines, we recently replicated the discriminative ability of a 9-SNP T1DGRS (developed in Europeans) in Indians but with lower power^17^.

In this study, we used an adequately powered and well-characterized case (T1D and T2D patients)-control cohort to investigate the possible reasons for the lower classification power of the GRS. We specifically focused on understanding the unique genetic architecture of Indians, reflecting significant differences in the risk allele frequencies of these genetic variants (both HLA and non-HLA) in Indians to ensure accurate risk assessment and improve diagnostic precision for T1D.

## Study Design and Methods

### Indian Cohorts

We included individuals of self-reported Indo-European ethnicity and classified them using specific criteria as described below.

### Type 1 diabetes (T1D)

The T1D cohort comprised 645 outpatients from the Diabetes Unit at KEMHRC, Pune, India. Patients were tested for glucose, C-peptide levels, islet autoantibodies (AA) including glutamic acid decarboxylase 65 (GAD65), insulinoma antigen 2 (IA2), and zinc transporter 8 (ZNT8). In addition to the clinical history, patients with random C-peptide concentrations below 600 pmol/L or positive for at least one AA within one year of diagnosis were included. Patients with an age at diagnosis less than 9 months and negative for AA were excluded. All T1D patients received insulin treatment from diagnosis (Supplementary Figure 1).

### Type 2 diabetes (T2D)

The T2D patients are from the WellGen study (n=1153) recruited at KEMHRC, Pune^18^. Briefly, T2D was defined based on the WHO1999 criteria, no history of ketoacidosis, response to oral hypoglycemic agents, and no insulin treatment for 5 years after diagnosis. Patients with possible exocrine pancreatic disease, monogenic diabetes etc. were excluded (n=37), leaving 1116 T2D patients. These patients were classified as T2D below 45 (n=788) and T2D above 45 (n=328) using age at diagnosis cut-off of 45 years.

### Controls

The controls were individuals without diabetes, verified using a 75g oral glucose tolerance test (WHO1999). The control group included parents of children from the Pune Children Study, which examined the relationship between a child’s birth weight and future risk of complex diseases^17,19^

### European Cohort

Summary statistics from the Type 1 Diabetes Genetics Consortium (T1DGC) cohort have been used in the study^20^.

### Measurement of C-peptide and islet autoantibodies in T1D patients

Serum samples were used for all the measurements. We estimated random C-peptide levels by direct electrochemiluminescence immunoassay (C-peptide kit, Roche Diagnostics GmBH, Germany; lower detection limit: 3.3 pmol/L; CV=0.6% at 33 pmol/L) on a Cobas e411 analyzer^21^. The islet autoantibodies were measured using a commercial ELISA kit (RSR Limited, Cardiff, UK). Patients were labelled positive using cut-off levels -GAD65 ≥ 5 IU/ml, IA2 ≥ 7.5 IU/ml and ZnT8 ≥ 15 IU/ml.

### High throughput genotyping and quality control analysis

We used Infinium Global Screening Array (GSA; Illumina Inc., USA) chips to generate genotype data on ∼6,50,000 SNPs in T1D patients and control participants as per manufacturer’s guidelines (GSA-v1=280 T1D patients and GSA-v3=317 T1D patients and 321 controls). Only overlapping SNPs between two chip versions were used for further analysis. After initial quality control (QC) using Genome Studio 2.0, a sample call rate of 95% and Gentrain >0.5, cluster separation >0.4 and AA/AB/BB Tdev < 0.06 were applied for the QC of SNP data. SNPs clustering poorly on Genome Studio were removed^22^ resulting in data on 585,848 SNPs from GSA-v1 in 280 T1D patients and 594,542 SNPs from GSA-v3 in 313 cases and 317 controls (Supplementary Figure 1). Both datasets were merged using the consensus call method in plink v1.9 resulting in 632,609 SNPs. Further QC was performed with filters, SNP call rate >95%, Hardy-Weinberg equilibrium p-value <10^-6^ and minor allele frequency >1%^23^. The final dataset for imputation included 369,275 SNPs from 593 T1D patients and 317 controls.

Genotype data on 1153 T2D patients was generated earlier using Affymetrix SNP 6.0 Chips (Affymetrix, CA, USA). After QC on 807,908 SNPs, using filters as described above, genotype data on 687,389 SNPs in 1116 T2D patients were used for further analysis.

### Genotype imputation

Genotype data from all the participants were prepared for imputation using willrayner toolkit. Genotype data on T2D patients were imputed separately as the data was generated on a different platform. Pre-phasing and imputation using the TOPMED R2 reference panel were performed with Eagle v2.4 and Minimac4 software respectively in the NIH imputation server^24,25^. We applied R^2^ ≥0.3 to select high-confidence imputation calls that retained 23,308,678 SNPs in T1D patients and controls each and 26,887,891 SNPs in T2D patients. HLA Genotype Imputation with Attribute Bagging (HIBAG) R package^26^ was employed to impute high-resolution four-digit classical HLA alleles using HIBAG prefit classifiers and genotypes specific to the HLA region. We used a GSA platform-specific prediction model, a multi-ethnic GSK-HLARES as the reference panel and a posterior probability cut-off ≥0.7 to ensure reliable imputed calls. The final dataset comprised 465 T1D patients (including 363 patients positive for at least one islet autoantibody) and 235 normal subjects. Phasing the HLA-DQ haplotypes (DQA1∼DQB1) was done using Bridging ImmunoGenomic Data Analysis Workflow Gaps R package^27^. The European data QC analysis and imputation are described in Sharp et al 2019^28^.

### Generation of genetic risk scores

The methods for GRS generation are described previously^28,29^. Due to low imputation accuracy for some SNPs included in the T1DGRS (R^2^ <0.3), we used proxy SNPs that are in complete linkage disequilibrium (r^2^=1 and D’=1 in the TOPMED data for SAS) with the missing SNPs. We generated three T1DGRSs comprising 9, 30 and 67 SNPs reported from European GWAS and compared their discriminative ability in Indian T1D patients versus T2D patients and normal subjects. T2D GRS was calculated with 330 SNPs due to missing data and lack of LD proxies on 8 SNPs, out of 338 independent hits selected from the largest multi-ancestry GWAS^30^.

### Statistical analyses

The discriminative ability of GRSs was evaluated by calculating the area under the curve (AUC) of the Receiver Operator Characteristic (ROC) curve and the confidence interval using the delong method^31^. The R packages, pROC and ROCit were used to estimate the ROC statistics and plot the curves^32^. A univariate logistic regression model was applied for haplotype association analysis using T1D status as the outcome variable and dosage of the haplotypes formed between HLA-DQA1 and HLA-DQB1, as the predictors. Youden’s index was derived from sensitivity and specificity as a performance metric. Z scores were calculated for T1DGRS and T2DGRS and used in robust diagnosis of T1D patients (Figure 4A). The multivariate regression model including T1D and T2D patients was employed to determine the predictive power of the combined use of T1DGRS and T2DGRS. Wilcoxon signed-rank test was used to compare T1DGRS and T2DGRS among diabetes groups. All statistical analyses were performed using R version 4.2.0.

## Results

### Clinical and demographic details

As per the criteria mentioned above, there were 593 T1D patients, 1116 T2D patients and 317 normal subjects. The gender distribution was largely similar in both patient groups. Close to 1/4th of T1D patients were negative for all three islet autoantibodies. Both autoantibody-negative and positive patients had similar age at diagnosis; however, the autoantibody-positive T1D patients had three-fold higher mean C-peptide levels than those negative for all islet autoantibodies (17.7 vs 6.2 pmol/L; P=0.067) but did not reach statistical significance (Supplementary Table 1).

### T1DGRS is discriminative of T1D in Indians but with lower ability than Europeans

Our previous study demonstrated that a 9-SNPs T1DGRS developed in Europeans, is discriminative of T1D vs T2D in Indians, albeit with lower discrimination by ROC AUC than Europeans^14^. On similar lines, the 9-SNPs T1DGRS discriminated T1D from T2D patients and controls (ROC-AUC (95%CI); 0.77 (0.75-0.79) and 0.77 (0.74-0.80) respectively) in the larger sample size in this study. The classification power using the 67 SNPs T1DGRS in T1D vs T2D [0.82 (0.80-0.85)] and T1D vs controls [0.83 (0.81-0.86)] was significantly higher compared to the 9-SNPs T1DGRS (P=1.28x10^-8^) and 30-SNPs T1DGRS (P=1.40x10^-4^). Hence, we conducted all further analyses with the 67-SNP T1DGRS (termed T1DGRS). Further, the density plot of T1DGRS, grouped based on diabetes types shows a clear separation of T1D from T2D patients (P=7.46x10^-108^) and controls (P=4.06x10^-61^) (Figure 1B) whereas T2D patients and controls had a similar distribution (P=0.86). However, T1DGRS-based discrimination of Indian T1D with non-T1D was still lower than the Europeans (0.92 vs 0.83, P<0.0001). We noted significant differences in the risk allele frequency at these variants and their effect size between Indians and Europeans. A statistically significant association with T1D was noted for 30 out of 67 SNPs and many showed a difference of 1.5 fold in the ORs (Supplementary Table 2). The remaining SNPs, though directionally consistent, did not replicate (p>0.05) in Indians. These observations strengthen our hypothesis of variable risk allele frequency and effect size as one of the reasons for the lower discriminatory potential of T1DGRS in Indians.

**Figure 1:**
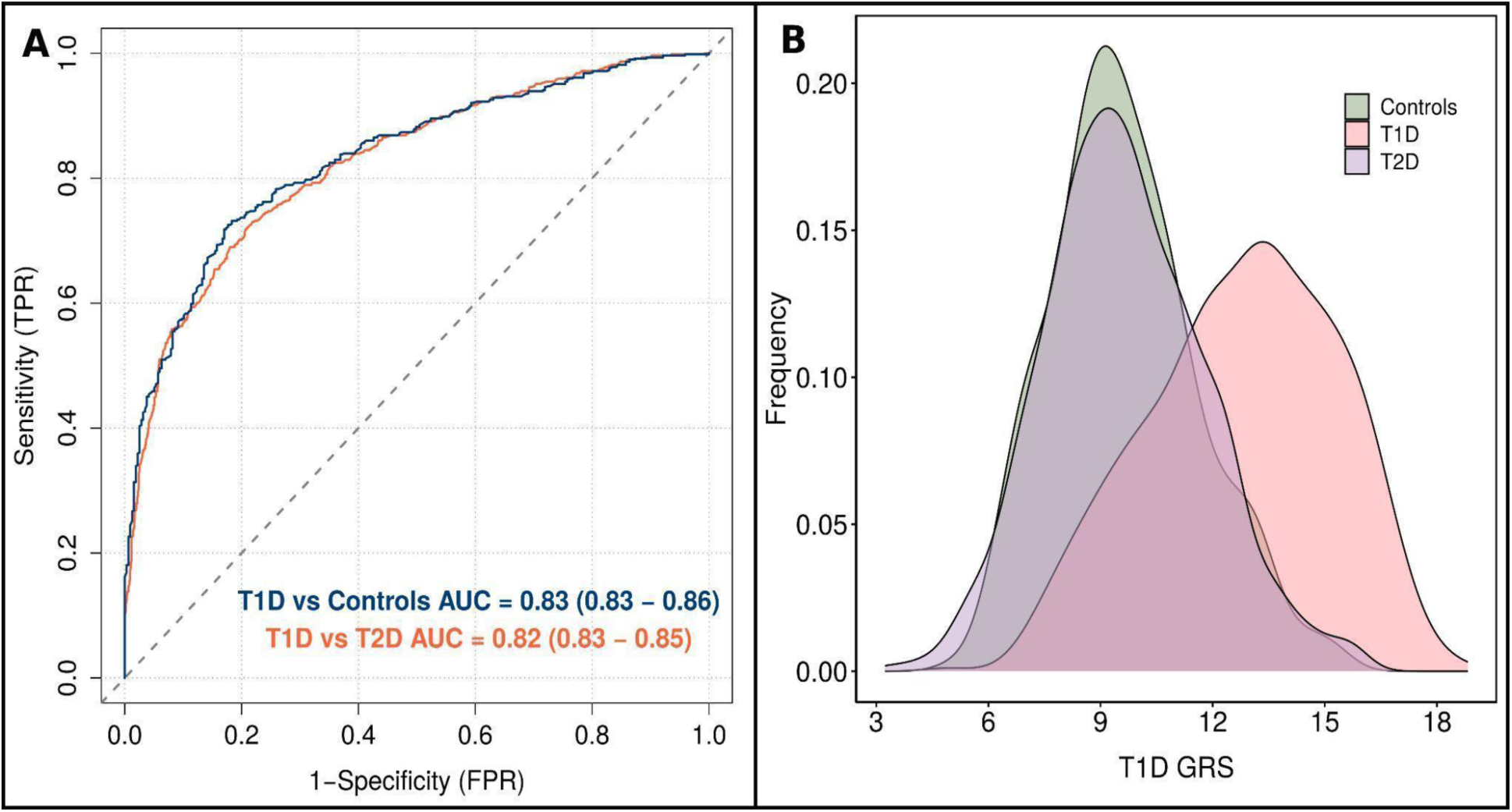
Discriminative ability of T1D genetic risk score in Indians. **A:** ROC curves indicating the discriminatory power of T1DGRS between T1D vs T2D and T1D vs Controls. **B:** Density plot showing how well T1DGRS separates T1D from T2D as well as controls. Colors indicate groups, the width represents the distribution and height indicates frequency. AUC, area under curve; ROC, receiver operating characteristic; T1D, type 1 diabetes; T2D, type 2 diabetes; FPR, false positive rate; TPR, true positive rate; GRS, genetic risk score.

### The frequency and effect size of T1D-associated HLA-DQ haplotypes are different in Indians

Out of 23 HLA-DQ haplotypes identified on 465 Indian T1D patients and 235 normal subjects, only 12 had a frequency ≥0.05%. Association analysis showed two haplotypes, DQA1*03:01∼DQB1*03:02 and DQA1*05:01∼DQB1*02:01 associated with T1D risk. The remaining ten are protective, which is directionally consistent with the Europeans (Figure 2A; Supplementary Table 3). While the majority of common haplotypes had comparable effect sizes and frequencies between the two ancestries, there were important differences. The most common protective HLA-DQ haplotype DQA1*01:02∼DQB1*06:02 present in 14% of Europeans, was detected in only 1.5% of Indians (Figure 2B). In contrast, another protective HLA-DQ haplotype, DQA1*01:03∼DQB1*06:01 was significantly more prevalent in Indians compared to Europeans (20.6% vs 0.4%), although the effect size was similar between the two populations. We noted that the effect size of the risk haplotype DQA1*05:01∼DQB1*02:01 in Indians was twice that in Europeans [7.78, 95%CI (5.29-11.83) vs 3.17 95%CI (2.99-3.37); P=1.4x10^-23^]. Further, it was inverse for the other risk haplotype DQA1*03:01∼DQB1*03:02, where Indians had significantly lower effect size than Europeans [2.52, 95%CI (1.76-3.68) vs 6.22, 95%CI (5.80-6.66); P=9.6x10^-7^]. Inclusion of DRB1 in the above two risk haplotypes, namely, DRB1*03:01∼DQA1*05:01∼DQB1*02:01 (DR3-DQ2) and the DRB1*04:01∼DQA1*03:01∼DQB1*03:02 (DR4-DQ8) also showed similar differences in the effect size between Indians and Europeans. Further, the T1D risk was several-fold higher for individuals compound heterozygous for this combination. We also noted that another protective haplotype DQA1*01:04∼DQB1*05:03 in the Europeans did not show any association in the Indians (P=0.94). Overall, these differences in the frequency and effect size of HLA-DQ haplotypes may account for the lower discriminative ability of T1DGRS in Indians compared to Europeans.

**Figure 2:**
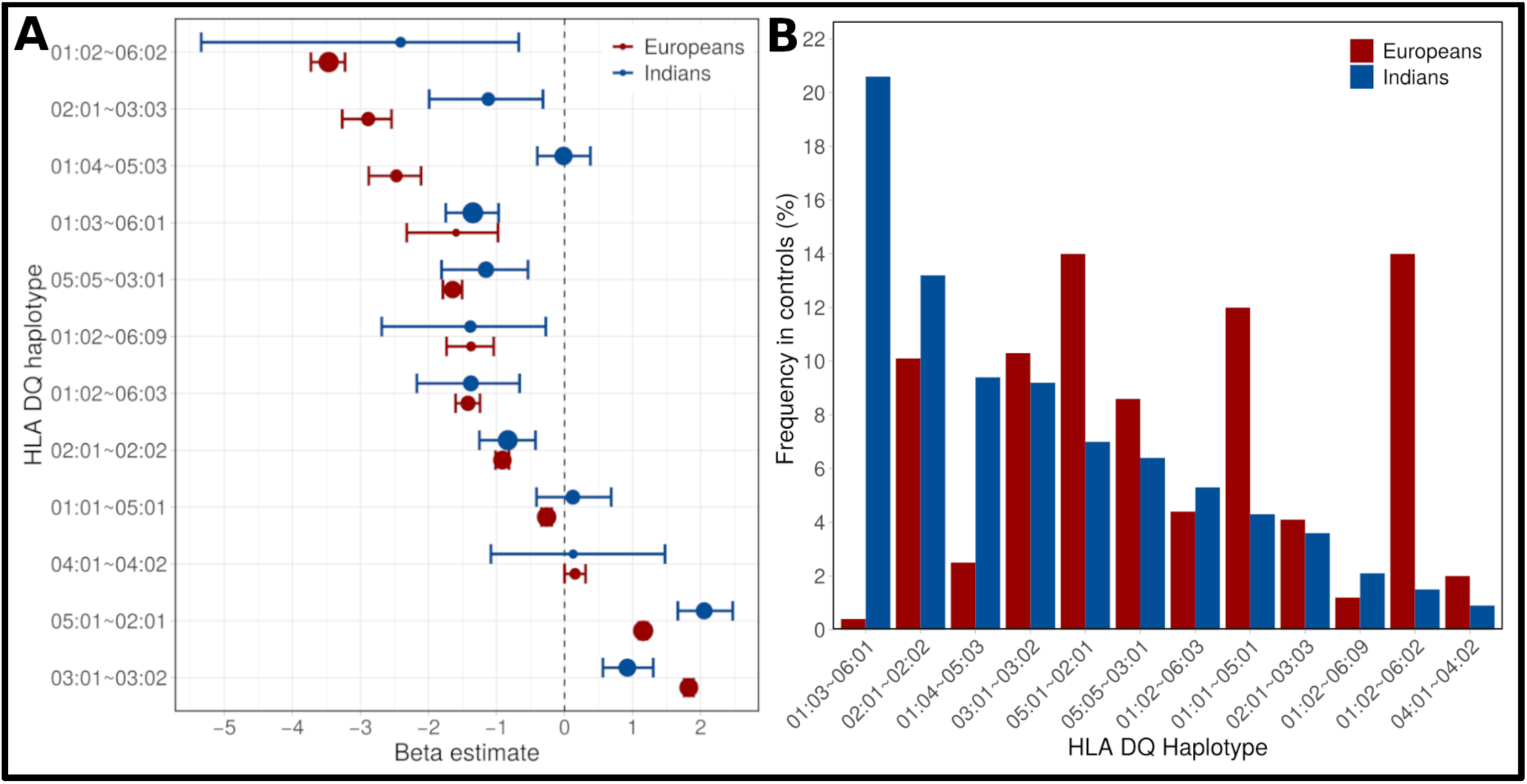
Comparison of frequency and the effect size of T1D-associated common HLA-DQ haplotypes between Indians and Europeans. **A:** Bar graph comparing the frequencies of 12 HLA-DQ haplotypes in Indian and European Controls; **B:** Forest plot showing differences in effect size distribution of 12 HLA-DQ haplotypes in Indians and Europeans. Circles and error bars represent the mean effect size (Beta estimate) and 95% confidence intervals respectively. The size of the circle represents the haplotype frequency in the controls. Indian T1D patients are considered only if at least one of the islet antibodies (GAD65, IA2, ZnT8) is positive.T1D, type 1 diabetes.

### The discriminative ability of T1DGRS in Indians is related to Islet autoantibody status and HLA SNPs

ROC-AUC analysis using T1DGRS showed better power to discriminate T1D patients positive for at least one islet autoantibody (T1D AA Pos; n=448) from T2D patients [0.85 (0.83-0.87)] (Figure 3A) and the controls [0.86 (0.83-0.88)], compared to all T1D patients taken together (Figure 1A). In contrast, the power of T1DGRS to separate T1D patients negative for all three autoantibodies (T1D AA Neg; n=145) from T2D patients and controls was significantly lower [(0.75 (0.70-0.80) and (0.70 (0.65-0.76)] respectively. Interestingly, the mean T1DGRS was higher for islet autoantibody-positive T1D patients compared to those negative for any islet autoantibody (13.0 vs 12.1; P=4.5x10^-4^). Similar results were obtained using the 9-SNP and 30-SNP T1DGRS with the above subgroup of T1D patients based on their islet autoantibody status (Supplementary Table 4). The inclusion of T2DGRS in the multivariate model did not improve the discriminatory power in either group, ruling out the possibility of autoantibody-negative T1D patients having T2D. These observations stress the important relationship between the polygenic score, islet autoantibody status and its discriminative ability.

**Figure 3:**
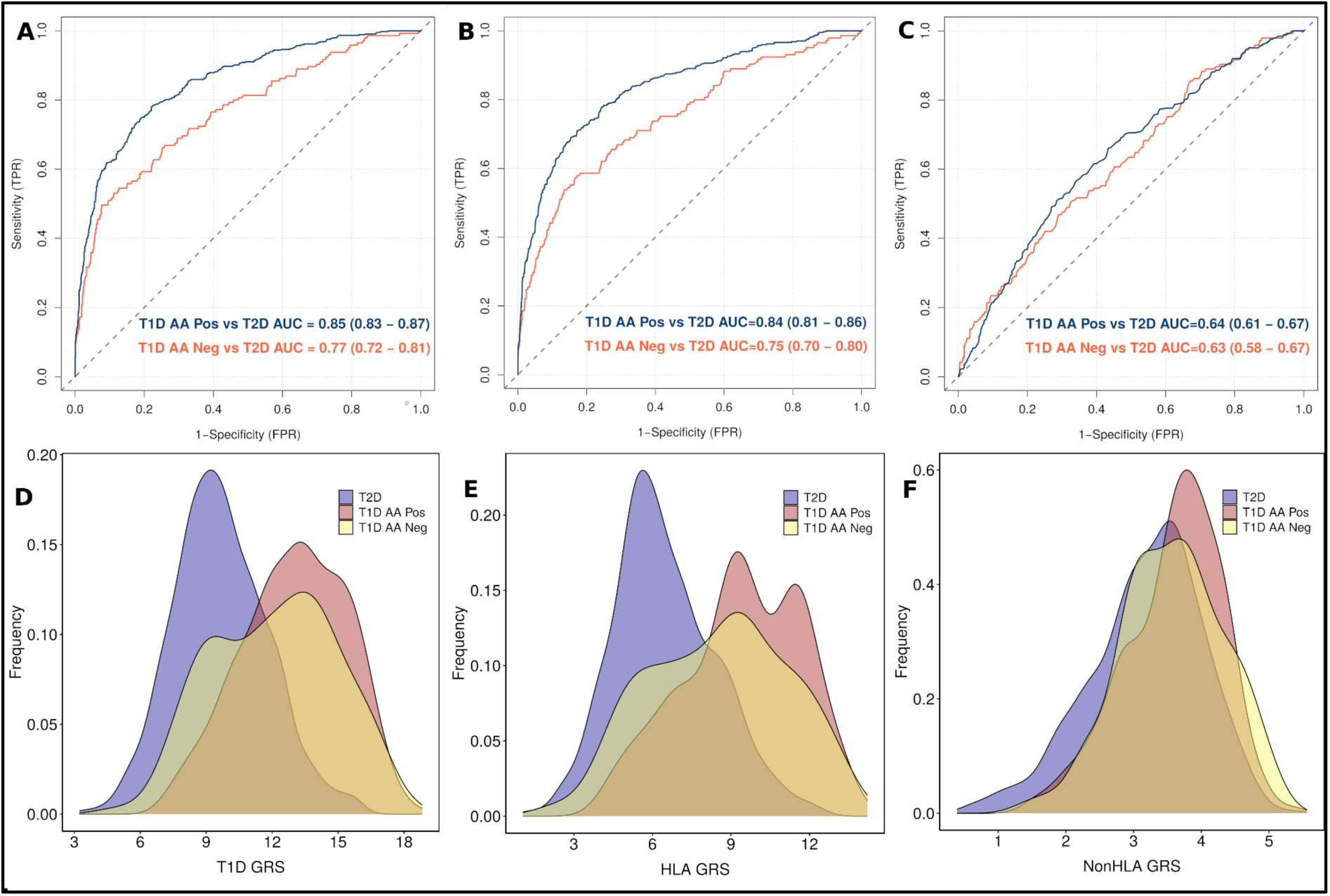
Discriminative ability of T1DGRS in Islet Autoantibody Positive (T1D AA Pos) and Autoantibody Negative (T1D AA Neg) patients. Autoantibody positive indicates positivity for at least one autoantibody (GAD65, IA2 and ZnT8) and those negative for all three autoantibodies were labeled as Autoantibody negative. T1DGRS was constructed by including **A:** all SNPs (n=67), **B:** Only HLA SNPs (n=35) and **C:** Only non-HLA SNPs (n=32). Density plot shows the discriminative ability using **D:** all SNPs, **E:** Only HLA SNPs and **F:** Only non-HLA SNPs. AUC, area under curve; T1D, type 1 diabetes; T2D, type 2 diabetes; FPR, false positive rate; TPR, true positive rate; GRS, genetic risk score.

**Figure 4:**
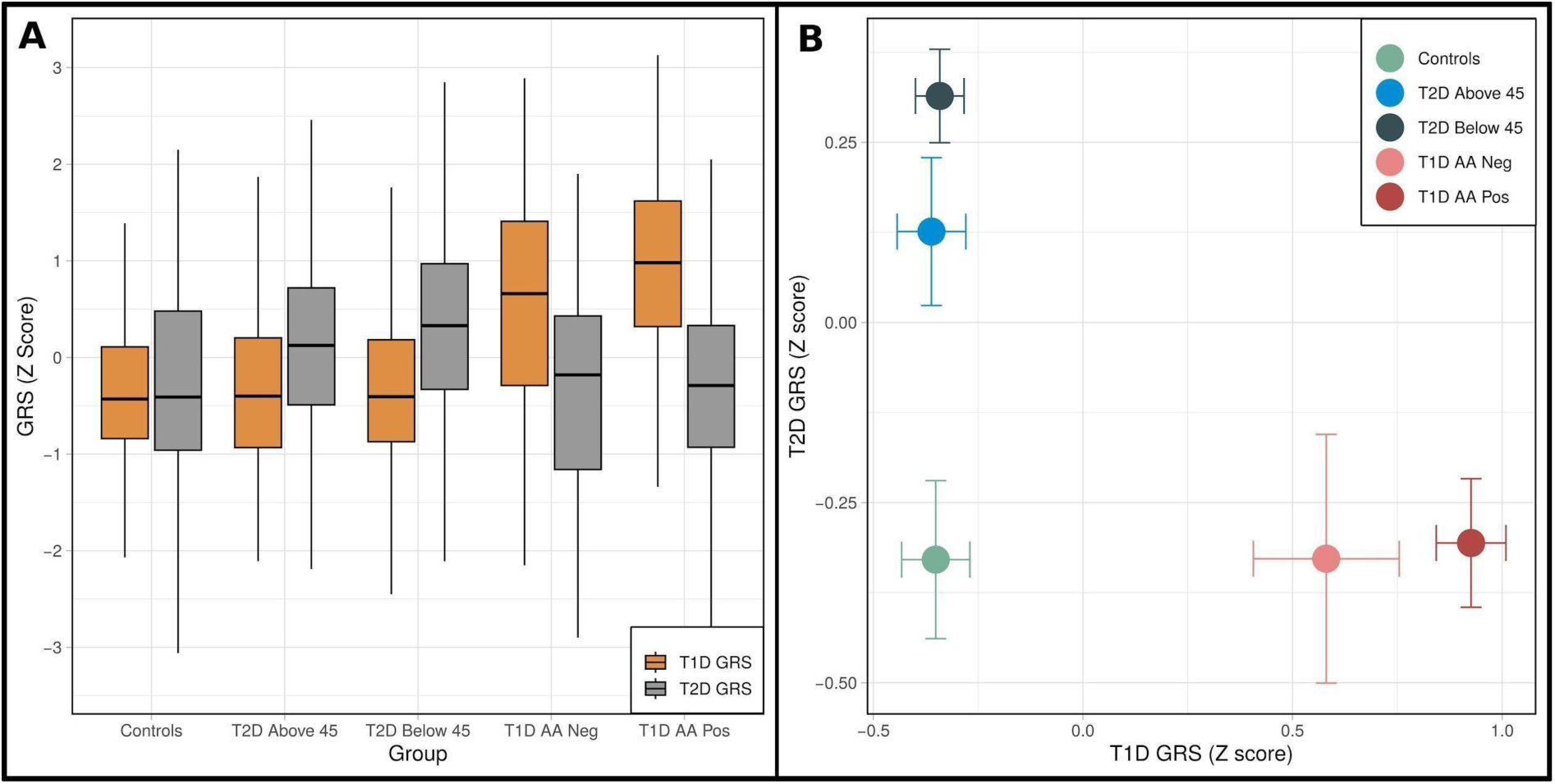
Differentiation of diabetes subtypes using T1D and T2DGRS (Z score). **A:** Box plots of T1D and T2DGRS (Z score) in two subtypes of diabetes. Data represented as median, IQR and whiskers represent the range (excluding outliers). **B:** T1D and T2DGRS plotted against each other to show the stratification of diabetes subtypes. Circles and error bars represent the Z-scored mean and 95% confidence intervals of T1DGRS (horizontal) and T2DGRS (vertical) respectively. Autoantibody positive (T1D AA Pos) indicates positivity for at least one autoantibody (GAD65, IA2 and ZnT8) and those negative for all three were labeled as Autoantibody negative (T1D AA Neg). T2D patients diagnosed before and after 45 years of age were denoted as T2D Below 45 and T2D Above 45 respectively. T1D, type 1 diabetes; T2D, type 2 diabetes; IQR, interquartile range; GRS, genetic risk score.

We further investigated the independent discriminative ability of HLA SNPs (HLAT1DGRS; n=35) (Figure 3B) and non-HLA SNPs (non-HLAT1DGRS; n=32) (Figure 3C) included in the T1DGRS. The classification power of HLA T1DGRS was significantly higher for T1D AA Pos than for T1D AA Neg with T2D patients [(0.83 (0.81-0.86) vs 0.73 (0.68-0.78); P=1.4x10^-4^] but remained similar for non-HLA T1DGRS [(0.64 (0.61-0.67) vs 0.63 (0.58-0.67); P=0.56)]. Similar results were noted between T1D patients and controls, based on patients’ islet autoantibody status (Supplementary Table 5). The T2DGRS had similar power in classifying T2D from T1D patients and non-diabetic controls [0.66 (0.63-0.69)] suggesting a limited utility of T2DGRS in robust discrimination (Supplementary Figure 2, Supplementary Table 6). However, keeping a cut-off of 45 years of age, the early onset T2D patients had higher T2DGRS thanthose with later onset (20.9 vs 20.8; P=1.7x10^-3^). Thus, two major types of diabetes can be discriminated by the independent use of T1DGRS (P=7.5x10^-108^) and T2DGRS (P=6.2x10^-27^) with different power; however, their combined use does not seem to provide any additional advantage.

To explore the utility of T1DGRS as an independent marker for robust diagnosis of T1D, we divided it into deciles in T1D patients and controls and calculated its sensitivity and specificity at each threshold (Supplementary Table 7). Using the median T1DGRS in controls (8.48) as the cut-off provides 52% sensitivity and 90% specificity. The best discriminative power was obtained at the T1DGRS of 11.37 which translates to the maximum Youden index of 0.59 and corresponds to 83% sensitivity and 75% specificity. The same in Europeans was noted at T1DGRS of 12.36 showing 89% sensitivity and 79% specificity^28^, indicating that population-specific thresholds are needed for the clinical utility of T1DGRS in other ancestries.

## Conclusions

It is known that phenotypic overlap between two major diabetes subtypes, T1D and T2D, especially in Indians, and the absence of sensitive and affordable markers creates uncertainties in robust diagnosis. This leads to clinical consequences as the optimal treatments differ for them. This study is an extension of our previous work showing the transferability of European-derived T1DGRS to discriminate T1D and T2D in Indians. We present four interesting observations from this study, 1) Using an adequately powered cohort and better T1DGRS comprising 67 SNPs, we demonstrate that the discriminative ability of the T1DGRS remains lower in Indians compared to the Europeans, 2) significant differences in the nature and distribution of HLA alleles/haplotypes and variability in their effect sizes exist between Indians and Europeans, which could contribute to the lower classification power of T1DGRS, 3) close to 1/4^th^ of Indian T1D patients are negative for islet autoantibody and HLA alleles in T1D patients correlate with their islet autoantibody status, and, 4) the discriminative ability of T1DGRS is lower in islet autoantibody negative T1D patients than those positive for any islet autoantibody, and these patients also have low T2D genetic risk suggesting the possibility of non-autoimmune pathogenesis or a non-classical T1D presentation. Combined, they may be responsible for the lower discriminatory potential of European-derived T1DGRS in Indians and raise important questions about the intersection of genetics, ethnicity, and health disparities in diabetes.

To the best of our knowledge, T1DGRS is the best predictive polygenic risk score marker ever shown for any disease or trait in Indians. Our results are consistent with other published studies suggesting that GRS portability from European studies may work well for South Asians compared to other ancestries such as East Asians or Africans^33^ but there are important differences between Indians and Europeans. It is worth noting that although better than the 9-SNP T1DGRS, the discriminative ability of the latest T1DGRS (using 67-SNPs) using a larger sample size continued to be lower in Indians. We found significant differences in risk allele frequencies and effect size of several T1D-associated SNPs between Indian and European subjects.

Variations in the HLA genes, especially DQA1 and DQB1 genes, are known to have a strong risk association with T1D and thus may have a big impact on the classification power of T1DGRS^28^. In contrast to 5 SNPs (17%) from the HLA region in the 30-SNP T1DGRS (the earlier GRS), the 67 SNP T1DGRS included 35 HLA SNPs (52%). Further, the haplotypes formed between HLA-DQA1 and HLA-DQB1 had the highest weightage in T1DGRS. We observed significant differences in HLA-DQ haplotype frequencies between Indians and Europeans, especially a significantly lower frequency of the strongest European T1D protective haplotype DQA1*01:02∼DQB1*06:02 in Indian controls and a significant variation in the effect size at two T1D risk haplotypes (DQA1*02:01∼DQB1*05:01 and DQA1*03:01∼DQB1*03:02) between Indians and Europeans. This further strengthens our earlier observation of a higher effect of DR3 than DR4 in Indians which is inverse in Europeans. Although the haplotypes are identified using the tag SNPs of DR3 and DR4, observations from the HLA haplotype analysis in this study are highly robust since we have used the four-digit haplotype in contrast to commonly used two-digit haplotype in all previous T1D studies from India. Recent reports demonstrate that four-digit HLA typing gives information on exact nucleotide variation and provides additional information that two-digit HLA typing lacks^34^.

It is difficult to speculate the exact reason for the differences in the nature of the association of individual haplotypes with T1D between Indians and Europeans; it may be surmised to be due to evolutionary dynamics shaping the survival of a population. Environmental disturbances are a major reason for the persistence of multiple alleles in a population, as no single allele confers protection against all diseases. Peculiarities of the Indian diet, gut microbiome and different genetic constitutions, e.g., nature and distribution of fucosyltransferase2 genetic variants further strengthen the argument^35^. These findings are also supported by other multi-ancestry studies showing that HLA haplotypes conferring T1D risk vary among the populations^36^. This evidence suggests that care should be taken in applying European-derived GRS in Indians and other non-European populations. This is further substantiated by the fact that although the latest T1DGRS score is reasonably similar in Indians and Europeans (11.37 vs 12.36 respectively), the Indian score has lower sensitivity and specificity than the Europeans. Thus, population-specific thresholds are needed for the clinical utilization of T1DGRS in other ancestries.

Our results demonstrate that T1DGRS has a good ability to discriminate between T1D and T2D patients. However, it may be difficult to diagnose T1D patients negative for all three islet autoantibodies correctly as their clinical phenotype overlaps with early-onset T2D. Consistent with the previous reports, the prevalence of islet autoantibody negative T1D patients was ∼25% compared to 5-10% in the Europeans^37,38^. This leaves a large chunk of T1D patients at risk of inaccurate diagnosis if only islet autoantibodies are used for Indian T1D patients. Thus T1DGRS is a good marker to facilitate the diagnosis as we show its utility in discrimination irrespective of the islet autoantibody status. Given the high prevalence of autoantibody-negative T1D patients and variability in genetic association (Supplementary Table 3), a genome-wide association study on a well-powered Indian cohort needs to be conducted.

## Strengths and Limitations

Our study has several strengths and few limitations. Patients included in the study were diagnosed using internationally accepted criteria and both cases and controls belonged to the same ethnicity (Indo-Europeans; one of the major population subgroups in India), confirming that results are unlikely to be affected by possible population substructure. One of the limitations could be the failure to estimate islet autoantibodies “At Diagnosis” for all T1D patients which might have overestimated the percentage of autoantibody negative patients since many autoantibody positive patients turn negative with treatment. However, the proportion of T1D patients negative for all three islet autoantibodies is comparable to other studies in India where autoantibodies were measured at diagnosis^38^.

In this study, we demonstrate the utility of T1DGRS in differentiating T1D patients from T2D patients. We also provide evidence of diversity and varying strength of association of both HLA alleles/haplotypes and several SNPs in non-HLA regions in Indians that is likely pivotal for the lower discriminative ability of T1DGRS in Indians. This study highlights an urgent need for further research to identify specific genetic risk variants and molecular pathways contributing to T1D and T2D across diverse populations that may provide more equitable and inclusive approaches to diabetes care.

## Additional Information

### Author Contributions

G.R.C., R.A.O., C.S.Y. and M.N.W. planned the study, provided the initial concept, study design and the analysis plan. C.S.Y. recruited the patients and P.K. coordinated the sample collection, measurement of biochemical data and proper data storage. M.S. generated the Indian genotype data under the supervision of G.R.C. A.S. processed genetic data, generated GRS, conducted all analyses and wrote the initial draft of the manuscript. D.F. conducted statistical analysis for the European samples. All authors reviewed the manuscript and agreed with the final version. G.R.C. is the guarantor of the work and takes responsibility for data integrity and accuracy of the data analysis.

### Ethics declaration

The collection of clinical data and use of bio-banked samples for the biochemical, immunological and genetic measurements was sanctioned by the Institutional Ethics Committee of the KEM Hospital Research Centre, Pune, India (KEMHRC ID No1737 & KEMHRC ID No PhD19) and all methods were performed following the relevant guidelines and regulations.

### Prior Presentation

A part of this research has been accepted for poster presentation at the American Society of Human Genetics (ASHG) Annual Meeting 2024.

## Data Availability

All data produced in the present study are available upon reasonable request to the authors

## Acknowledgements

We thank the participants in the study. The funding from Diabetes UK, London, UK and the Council of Scientific and Industrial Research (CSIR), Ministry of Science and Technology, Government of India, New Delhi, India is gratefully acknowledged. We also acknowledge the help of Mr Inder Deo Mali from CSIR-CCMB in isolating high-quality genomic DNA from blood samples in various cohorts.

---

**Supplementary Figure 1:**
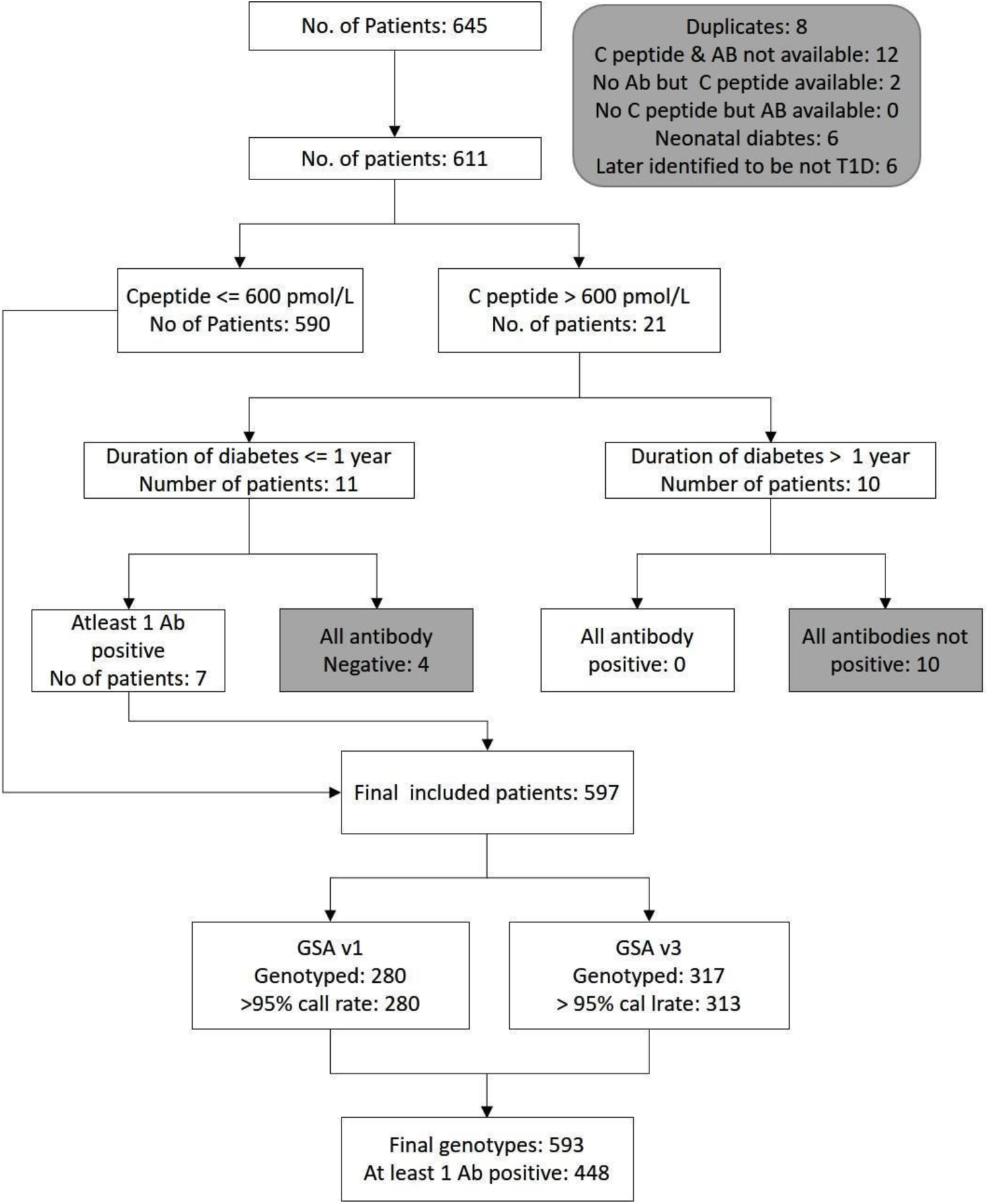
Consort diagram representing the type 1 diabetes patients inclusion/exclusion criteria. Sample removal steps are highlighted in gray. Ab, Autoantibody

**Supplementary Figure 2:**
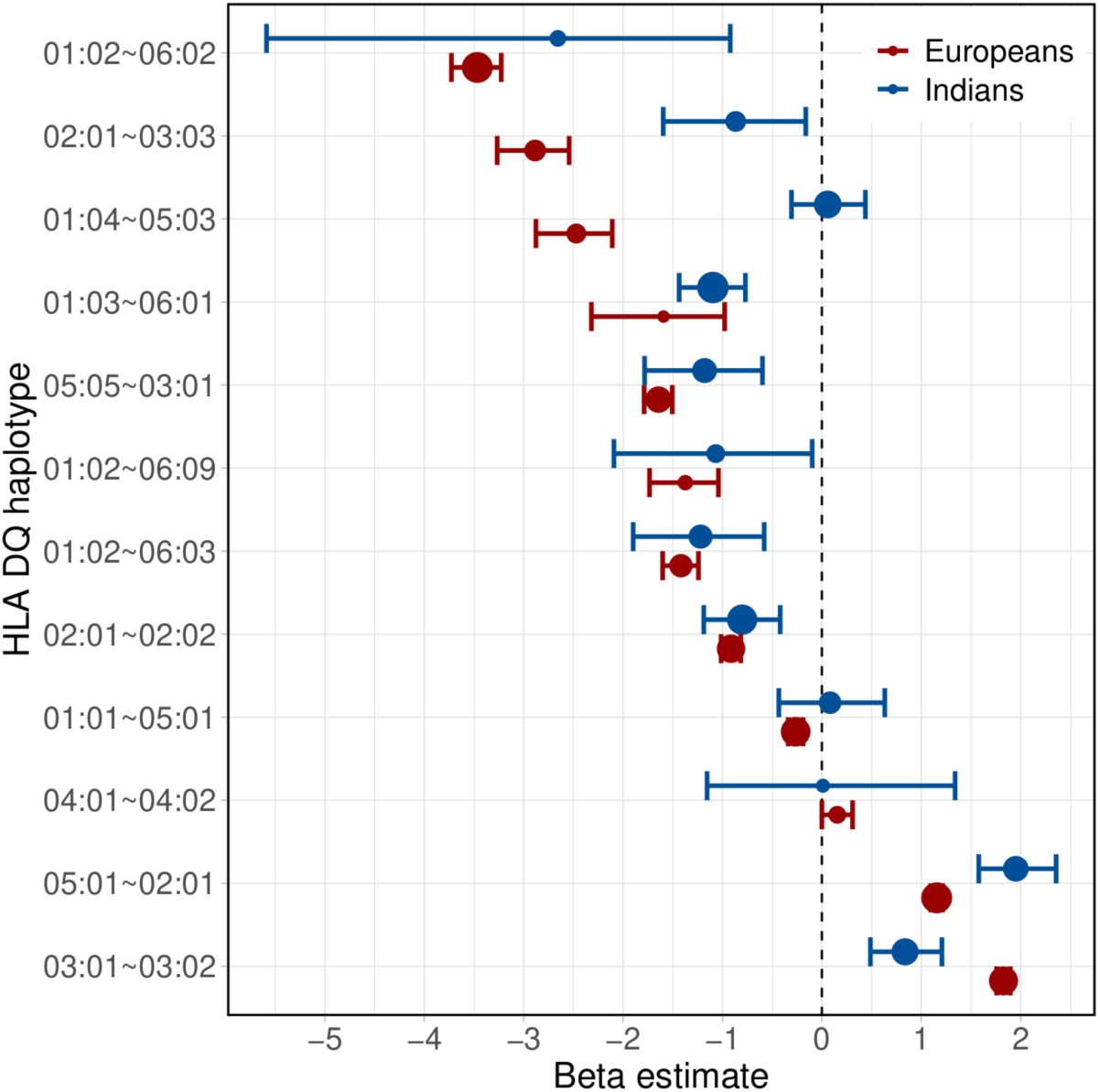
Forest plot showing differences in the effect size distribution of common HLA-DQ haplotypes between Indian and European T1D patients. Circles and error bars represent the mean effect size (Beta estimate) and 95% confidence intervals respectively. The size of the circle represents the haplotype frequency in the controls. Only T1D patients positive for at least one of the islet autoantibodies (GAD65, IA2, ZnT8) were. T1D, type 1 diabetes.

**Supplementary Figure 3:**
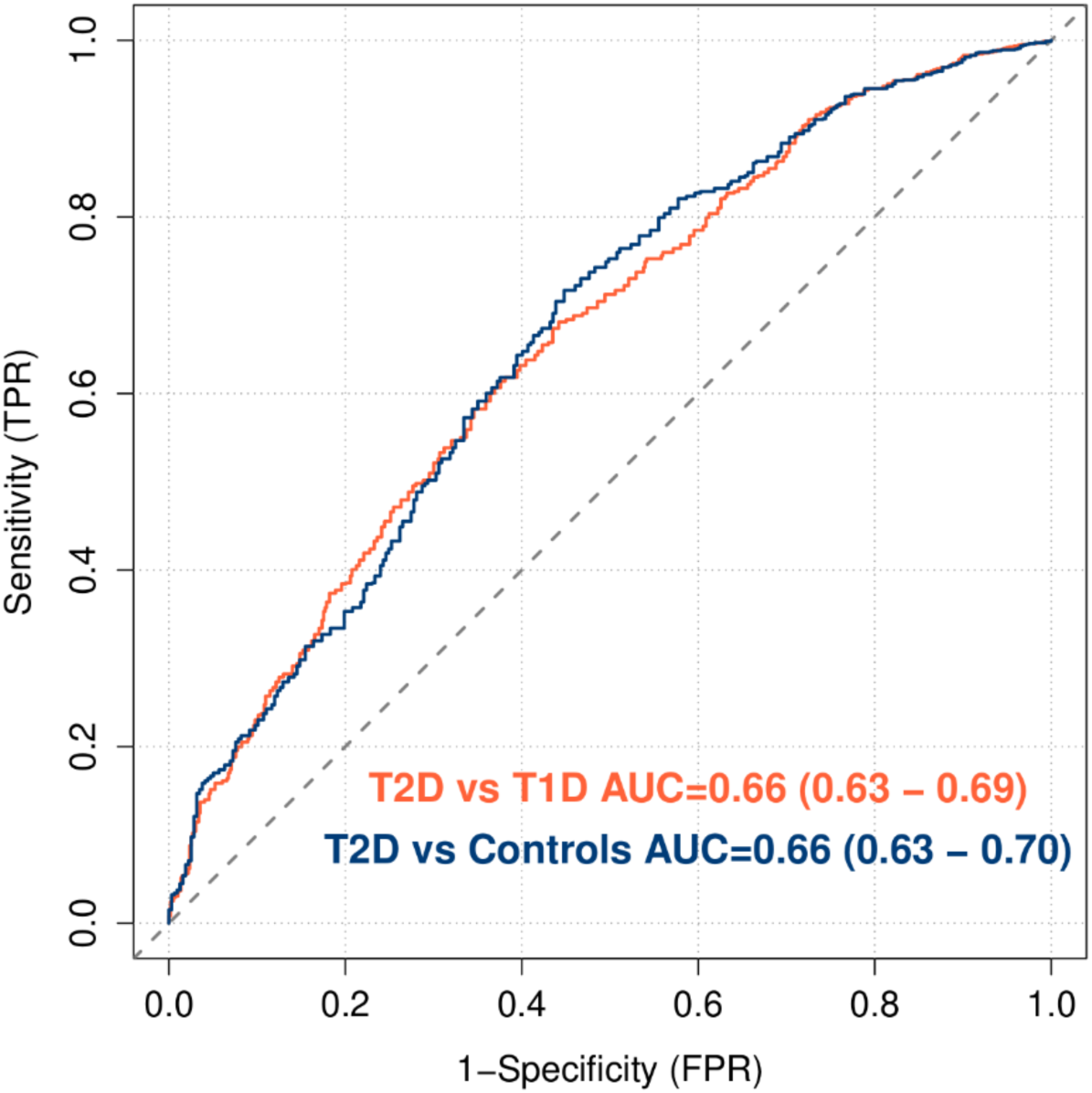
ROC curves indicating the power of T2DGRS in discriminating between T2D vs T1D and T2D vs Controls. AUC, area under curve; T1D, type 1 diabetes; T2D, type 2 diabetes; ROC, receiver operating characteristic; FPR, false positive rate; TPR, true positive rate; GRS, genetic risk score.

## References

1. Magliano D, Boyko EJ. IDF Diabetes Atlas. 10th edition. International Diabetes Federation; 2021.

2. The Expert Committee on the Diagnosis and Classification of Diabetes Mellitus*. Follow-up Report on the Diagnosis of Diabetes Mellitus. Diabetes Care. 2003;26(11):3160–3167. doi:10.2337/diacare.26.11.3160

3. Thomas NJ, Jones SE, Weedon MN, Shields BM, Oram RA, Hattersley AT. Frequency and phenotype of type 1 diabetes in the first six decades of life: a cross-sectional, genetically stratified survival analysis from UK Biobank. Lancet Diabetes Endocrinol. 2018;6(2):122–129. doi:10.1016/S2213-8587(17)30362-5

4. Gregory GA, Robinson TIG, Linklater SE, et al. Global incidence, prevalence, and mortality of type 1 diabetes in 2021 with projection to 2040: a modelling study.

5. Hills AP, Arena R, Khunti K, et al. Epidemiology and determinants of type 2 diabetes in south Asia. Lancet Diabetes Endocrinol. 2018;6(12):966–978. doi:10.1016/S2213-8587(18)30204-3

6. Thomas NJ, Lynam AL, Hill AV, et al. Type 1 diabetes defined by severe insulin deficiency occurs after 30 years of age and is commonly treated as type 2 diabetes. Diabetologia. 2019;62(7):1167–1172. doi:10.1007/s00125-019-4863-8

7. Muñoz C, Floreen A, Garey C, et al. Misdiagnosis and Diabetic Ketoacidosis at Diagnosis of Type 1 Diabetes: Patient and Caregiver Perspectives. Clin Diabetes. 2019;37(3):276–281. doi:10.2337/cd18-0088

8. Yki-Järvinen H. Combination Therapies With Insulin in Type 2 Diabetes. Diabetes Care. 2001;24(4):758–767. doi:10.2337/diacare.24.4.758

9. Sims EK, Besser REJ, Dayan C, et al. Screening for Type 1 Diabetes in the General Population: A Status Report and Perspective. Diabetes. 2022;71(4):610–623. doi:10.2337/dbi20-0054

10. Bingley PJ. Clinical Applications of Diabetes Antibody Testing. J Clin Endocrinol Metab. 2010;95(1):25–33. doi:10.1210/jc.2009-1365

11. Barrett JC, Clayton DG, Concannon P, et al. Genome-wide association study and meta-analysis find that over 40 loci affect risk of type 1 diabetes. Nat Genet. 2009;41(6):703–707. doi:10.1038/ng.381

12. Robertson CC, Inshaw JRJ, Onengut-Gumuscu S, et al. Fine-mapping, trans-ancestral and genomic analyses identify causal variants, cells, genes and drug targets for type 1 diabetes. Nat Genet. 2021;53(7):962–971. doi:10.1038/s41588-021-00880-5

13. Chiou J, Geusz RJ, Okino ML, et al. Interpreting type 1 diabetes risk with genetics and single-cell epigenomics. Nature. 2021;594(7863):398–402. doi:10.1038/s41586-021-03552-w

14. Fitipaldi H, Franks PW. Ethnic, gender and other sociodemographic biases in genome-wide association studies for the most burdensome non-communicable diseases: 2005–2022. Hum Mol Genet. 2023;32(3):520–532. doi:10.1093/hmg/ddac245

15. Chandak GR, Idris MM, Reddy DN, et al. Absence of PRSS1 mutations and association of SPINK1 trypsin inhibitor mutations in hereditary and non-hereditary chronic pancreatitis. Gut. 2004;53(5):723. doi:10.1136/gut.2003.026526

16. Yajnik CS, Janipalli CS, Bhaskar S, et al. FTO gene variants are strongly associated with type 2 diabetes in South Asian Indians. Diabetologia. 2009;52(2):247–252. doi:10.1007/s00125-008-1186-6

17. Harrison JW, Tallapragada DSP, Baptist A, et al. Type 1 diabetes genetic risk score is discriminative of diabetes in non-Europeans: evidence from a study in India. Sci Rep. 2020;10(1):9450. doi:10.1038/s41598-020-65317-1

18. Chandak GR, Janipalli CS, Bhaskar S, et al. Common variants in the TCF7L2 gene are strongly associated with type 2 diabetes mellitus in the Indian population. Diabetologia. 2007;50(1):63–67. doi:10.1007/s00125-006-0502-2

19. Yajnik CS, Fall CHD, Vaidya U, et al. Fetal Growth and Glucose and Insulin Metabolism in Four-year-old Indian Children. Diabet Med. 1995;12(4):330–336. doi:10.1111/j.1464-5491.1995.tb00487.x

20. RICH SS, CONCANNON P, ERLICH H, et al. The Type 1 Diabetes Genetics Consortium. Ann N Y Acad Sci. 2006;1079(1):1–8. doi:10.1196/annals.1375.001

21. Oram RA, Jones AG, Besser REJ, et al. The majority of patients with long-duration type 1 diabetes are insulin microsecretors and have functioning beta cells. Diabetologia. 2014;57(1):187–191. doi:10.1007/s00125-013-3067-x

22. Guo Y, He J, Zhao S, et al. Illumina human exome genotyping array clustering and quality control. Nat Protoc. 2014;9(11):2643–2662. doi:10.1038/nprot.2014.174

23. Purcell S, Neale B, Todd-Brown K, et al. PLINK: A Tool Set for Whole-Genome Association and Population-Based Linkage Analyses. Am J Hum Genet. 2007;81(3):559–575. doi:10.1086/519795

24. Das S, Forer L, Schönherr S, et al. Next-generation genotype imputation service and methods. Nat Genet. 2016;48(10):1284–1287. doi:10.1038/ng.3656

25. Taliun D, Harris DN, Kessler MD, et al. Sequencing of 53,831 diverse genomes from the NHLBI TOPMed Program. Nature. 2021;590(7845):290–299. doi:10.1038/s41586-021-03205-y

26. Zheng X, Shen J, Cox C, et al. HIBAG—HLA genotype imputation with attribute bagging. Pharmacogenomics J. 2014;14(2):192–200. doi:10.1038/tpj.2013.18

27. Pappas DJ, Marin W, Hollenbach JA, Mack SJ. Bridging ImmunoGenomic Data Analysis Workflow Gaps (BIGDAWG): An integrated case-control analysis pipeline. Spec Issue Histocompat Immunogenetics Data Resour. 2016;77(3):283–287. doi:10.1016/j.humimm.2015.12.006

28. Sharp SA, Rich SS, Wood AR, et al. Development and Standardization of an Improved Type 1 Diabetes Genetic Risk Score for Use in Newborn Screening and Incident Diagnosis. Diabetes Care. 2019;42(2):200–207. doi:10.2337/dc18-1785

29. Oram RA, Patel K, Hill A, et al. A Type 1 Diabetes Genetic Risk Score Can Aid Discrimination Between Type 1 and Type 2 Diabetes in Young Adults. Diabetes Care. 2015;39(3):337–344. doi:10.2337/dc15-1111

30. Mahajan A, Spracklen CN, Zhang W, et al. Multi-ancestry genetic study of type 2 diabetes highlights the power of diverse populations for discovery and translation. Nat Genet. 2022;54(5):560–572. doi:10.1038/s41588-022-01058-3

31. DeLong ER, DeLong DM, Clarke-Pearson DL. Comparing the areas under two or more correlated receiver operating characteristic curves: a nonparametric approach. Biometrics. 1988;44(3):837–845.

32. Robin X, Turck N, Hainard A, et al. pROC: an open-source package for R and S+ to analyze and compare ROC curves. BMC Bioinformatics. 2011;12(1):77. doi:10.1186/1471-2105-12-77

33. Ding Y, Hou K, Xu Z, et al. Polygenic scoring accuracy varies across the genetic ancestry continuum. Nature. 2023;618(7966):774-781. doi:10.1038/s41586-023-06079-4

34. Huang Y, Dinh A, Heron S, et al. Assessing the utilization of high-resolution 2-field HLA typing in solid organ transplantation. Am J Transplant. 2019;19(7):1955–1963. doi:10.1111/ajt.15258

35. Nongmaithem SS, Joglekar CV, Krishnaveni GV, et al. GWAS identifies population-specific new regulatory variants in FUT6 associated with plasma B12 concentrations in Indians. Hum Mol Genet. 2017;26(13):2551–2564. doi:10.1093/hmg/ddx071

36. Michalek DA, Tern C, Zhou W, et al. A multi-ancestry genome-wide association study in type 1 diabetes. Hum Mol Genet. 2024;33(11):958–968. doi:10.1093/hmg/ddae024

37. Basu M, Pandit K, Banerjee M, Mondal SA, Mukhopadhyay P, Ghosh S. Profile of Auto-antibodies (Disease Related and Other) in Children with Type 1 Diabetes. Indian J Endocrinol Metab. 2020;24(3). https://journals.lww.com/indjem/fulltext/2020/24030/profile_of_auto_antibodiesdisease_related_and.6.aspx

38. Vipin VP, Zaidi G, Watson K, et al. High prevalence of idiopathic (islet antibody-negative) type 1 diabetes among Indian children and adolescents. Pediatr Diabetes. 2021;22(1):47–51. doi:10.1111/pedi.13066

